# Non-Contact Optical Blood Pressure Biometry Using AI-Based Analysis of Non-Mydriatic Fundus Imaging

**DOI:** 10.1101/2025.01.06.25320084

**Authors:** Idan Bressler, Dolev Dollberg, Rachelle Aviv, Danny Margalit, Alon Harris, Brent Siesky, Tsontcho Ianchulev, Zack Dvey-Aharon

## Abstract

**Background:** This study was developed to determine whether a machine learning model could be developed to assess blood pressure with accuracy comparable to arm cuff measurements.

**Methods:** A deep learning model was developed based on the UK Biobank dataset and was trained to detect both systolic and diastolic pressure. The hypothesis was formulated after data collection and before the development of the model. Comparison was conducted between arm cuff measurements, as ground truth, and results from the model, using Mean Absolute Error, Mean Squared Error, and Coefficient of Determination (R^2).

**Results:** Systolic pressure was measured with 9.81 Mean Absolute Error, 165.13 Mean Squared Error and 0.36 R^2. Diastolic pressure was measured with 6.00 Mean Absolute Error, 58.21 and 0.30 R^2.

**Conclusions:** This model improves on existing research and shows errors comparable to the variability of hand cuff measurements. The use of fundus images to assess blood pressure may be more indicative of long-term hypertension. Additional trials in clinical settings may be necessary, as well as additional prospective studies to validate results.

## Introduction

Blood pressure (BP) measurement is one of the most common key health assessment procedures utilized across various medical settings.^1^ Elevated BP, or hypertension, is a leading cause for cardiovascular disease, affecting approximately 30% of the adult population worldwide.^2^ Hypertension further causes or accelerates disease in multiple additional systems, including in renal, eye, and brain tissues.^3,4^ Elevated BP is also a significant driver of disease disparities in persons of African descent, contributing to disproportionately elevated rates of stroke and cardiovascular disease.^5^ A large and growing prevalence of global hypertension and an increase in cardiovascular disease burden necessitates identifying novel, highly accessible approaches for improving BP diagnostics and lowering the impact from associated diseases.

Hypertension can be defined by a persistent BP of 140/90 mm Hg (systolic/diastolic),^6^ whereas systolic hypertension involves only systolic BP ≥ 140 and diastolic hypertension is defined as only diastolic BP ≥ 90. Hypertension can be further subdivided into grade one (140-159/90-99 mmHg) and grade two hypertension (160/100 mmHg); the latter requiring immediate drug treatment in all patients. The US uses slightly different definitions with hypertension starting with 130/80 mmHG.^7^ Isolated systolic hypertension and an increase in pulse pressure is frequently observed in elderly populations due to stiffening of arteries while it is also the most common form of essential hypertension found in young adults.^8^

Management of hypertension is focused on reducing BP levels to within normal range through changes in lifestyle and diet, and with targeted medications.^9^ BP measurement is typically done while the patient is sitting or supine using standard or ambulatory arm cuff devices.^10^ It is important to note, however, that cuff BP measurements have shown significant intrapatient variability of up to 10 mmHg systolic BP on consecutive readings.^11,12^ In addition, patients undergoing cuff BP measurements are known to have frequently elevated BP readings when suffering from the often stress induced “white coat” hypertension phenomenon.^13^

Hypertension can manifest pathology in the retina in various ways, with notable long term changes occurring in the microvascular structure, optic disc, and other retinal features.^14^ Collectively these changes are referred to as “hypertensive retinopathy,” and if left untreated, may cause vision loss.^15^ Mechanistically, hypertension initially causes retinal arterioles to undergo narrowing due to local autoregulatory mechanisms. Persistently elevated pressures lead to changes in the vessel walls, which manifest as severe generalized and focal arteriolar narrowing, arteriovenous (AV) nicking, and opacification of vessels walls, termed silver or copper wiring.^14,15^ Severe BP elevation can also disturb the blood retinal barrier and cause hemorrhages, exudation, ischemia, and disc edema. Studies have found that AV nicking and generalized narrowing represent effects of chronic hypertension.^16,17^

Hypertensive retinopathy is a cornerstone of eye exams and large epidemiologic studies have reported the association of retinopathy signs with hypertensive end-organ microvascular damage in conditions such as stroke, dementia, and coronary heart disease. The estimated prevalence of hypertensive retinopathy among patients without diabetes aged 40 years or older varies from 2-14% in large population-based studies.^18^ One study, based on the Beaver Dam Eye Study population, found a 9.2% incidence of retinopathy in hypertensive patients.^19^ In addition to retinopathy, hypertension has been identified as a potential contributor to the glaucomatous disease process.^4^ In persons of African descent, both elevated rates of hypertension and directly assessed reductions in retinal vascular densities and retinal blood flow have been linked to glaucoma progression.^4,20^

The retina is unique as the only body tissue where the micro-vasculature can be directly imaged, observed, and quantified via biomarkers using non-invasive optics.^4^ The high accessibility of retinal imaging has also established it as a source of data for systemic diseases including diabetes and hypertension. Examination of the retinal microvasculature in the context of hypertension may provide evidence of long-term hypertensive induced vascular damage.

This is in contrast with cuff BP measurements, which provide only a snapshot of arm pressure at the time of the measurement. Acute cuff BP readings are further complicated by the fact that systolic and diastolic BP exhibit a diurnal pattern, and readings can be significantly affected by the time of day in which they are assessed. This leads to periods of chronic unobserved hypertension or hypotension that are not observed or accounted for during normal outpatient medical visits. Cuff BP measurements therefore have high variation due to diurnal flux, the specific BP procedures and device used, and other extrinsic and behavioral factors.^1,21^

Although much has been studied on the predictive value of retina micro-vascular biomarkers for chronic compound damage over time, non-invasive touchless optical retinal imaging has not been previously used to predict and estimate systemic BP as a surrogate to sphygmomanometric BP measurements using conventional algorithmic methodologies. More recently, artificial intelligence (AI) models have been deployed to investigate potential clinical utility,^22^ as the quest for cuffless BP measurements is gaining momentum in the scientific and medical communities.^23^ Previous work has demonstrated the ability to detect BP from fundus images,^22^ and is suggestive of the ability to detect retinal structure changes according to the patient’s BP before detectable hypertensive retinopathy changes are present.

Due to the complexity of assessing accurate BP and the vast amount of data sourced from retinal tissues, standard statistical approaches have often failed in discovery. More recently, AI methods for automatic assessment of the fundus using deep learning (DL) have shown promising results in detection of various conditions.^24^ Specifically, autonomous DR screening has received FDA approval,^25^ opening the door to the implementation of further similar applications in a clinical setting, such as BP measurement. In this analysis, we aim to assess systemic BP from fundus images using a DL model and to examine its accuracy compared to traditional cuff BP measurement.

## Methods

### Data

This was a retrospective analysis utilizing the UK-BioBank dataset,^26^ which consists of 105,564 45° angle, macula-centered fundus photography images from 51,791 patients. The dataset consists of two images per patient per visit, one from each eye. This research has been conducted using the UK Biobank Resource under Application Number 75750.

At the imaging visit patients underwent additional tests, including two automatic arm cuff BP assessments, taken at the same visit, for both systolic and diastolic BP measurements; the readings were taken at a short interval, and the second record was selected as the ground truth in order to reduce the “white coat” effect. Although cuff BP measurements have the limitations discussed earlier, they have been the prevailing standard of care.

The data selected for this study excluded any patient with a record of BP medication, as the aim of this analysis is to reveal the long-term effects of hypertension. As such, for these patients, there may be a large gap between their current measured BP and the state of the retina. The resulting dataset consisted of 105,564 images from 51,791 patients (with some patients having more than one visit). Additional demographic data is presented in **table 1**.

**Table 1:** population statistics.

| Field | Images | Patients | Mean Age (SD) | Gender (% Female) | Ethnicity (fraction) | Blood Pressure |
| --- | --- | --- | --- | --- | --- | --- |
| Value | 105,564 | 51,791 | 55.95 (8.31) | 59 | British (Unspecified) = 0.84<br>Non-British = 0.16<br>Non-British Ethnicities (fraction of group):<br>White = 0.51<br>Indian = 0.10<br>Caribbean = 0.08<br>East Asian = 0.06<br>African = 0.05<br>Other = 0.2 | Systolic (mmHG): 129 (16)<br>Diastolic (mmHG): 79 (9) |

### Pre-Processing

Image pre-processing was performed in two steps. Firstly, the images were resized to a standard size, and secondly the image background was cut along the convex hull which contains the circular border between the image and the background.

### Quality Assessment

A heuristic tool for image quality assessment was developed based on detecting visibility of fundus specific characteristics. The given quality score for an image was an aggregation of the visibility score from multiple areas within the fundus image.

### Model Training

Following image quality assessment, 14,709 images (∼14% of the data) were filtered, leaving 90,855 images for analysis. The data was then divided into training, validation, and test datasets consisting of 80%, 10%, and 10% of the data respectively. Customary morphologic and color augmentation was applied during training.

A regression neural network with two outputs (systolic and diastolic values) was trained. The model used was a convolutional neural network (CNN). The model architecture was fitted to best balance the model performance vs. model complexity tradeoff, ending with a model of ∼28 million parameters and a size of ∼110 MB. Hyperparameter tuning was done over the validation set. The results from both eyes were combined to provide an overall patient BP reading.

### Statistical Analysis

The metrics used for assessment were Mean Absolute Error (MAE), Mean Squared Error (MSE), and Coefficient of Determination (R^2). For each metric the bias corrected and accelerated bootstrap method^27^ was used to produce a 95% confidence interval.

### Model Visualization

The Grad-CAM method^28^ was used to visualize the model areas of interest in the form of a heatmap, as shown in **Figure 1**.

**Figure 1:**
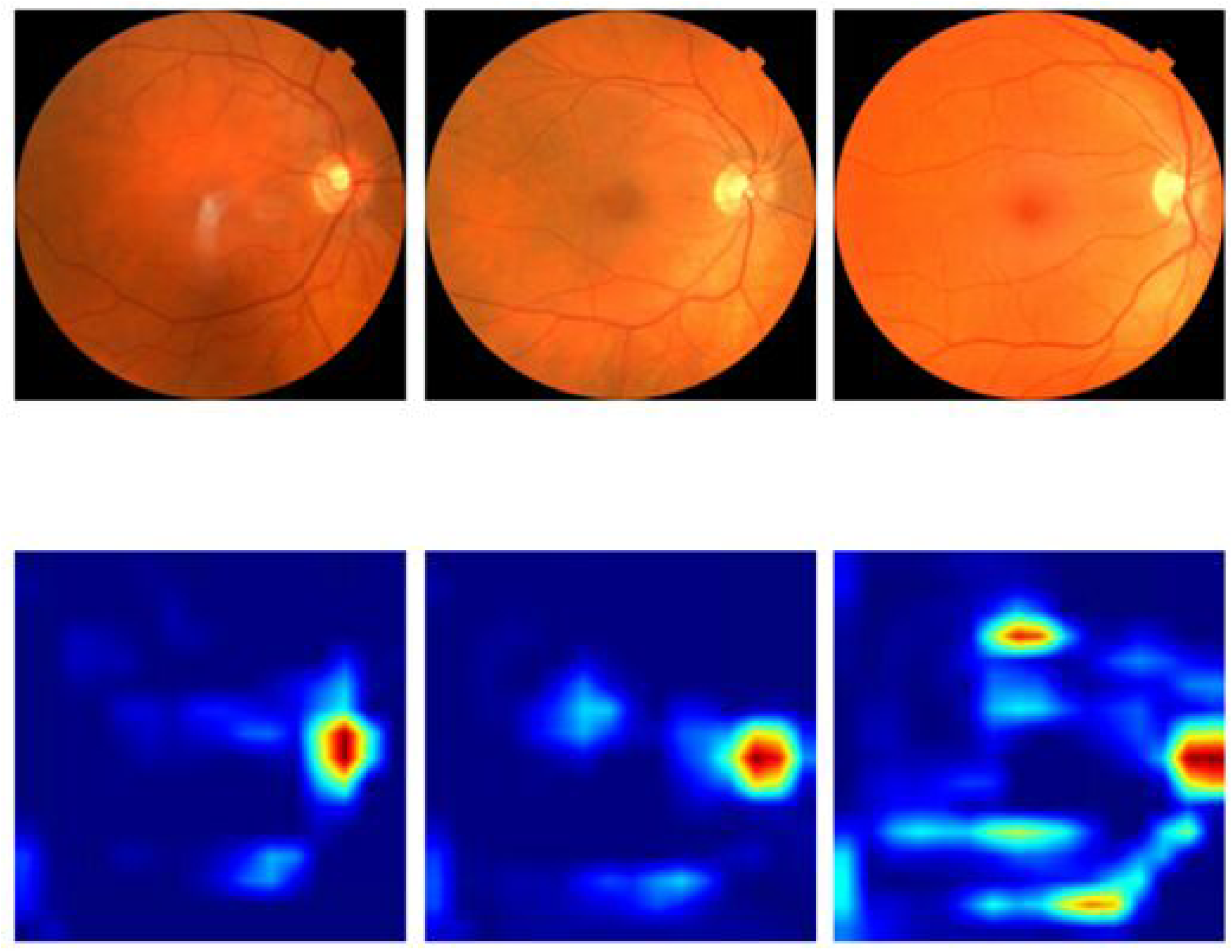
Grad-CAM results for 3 right eyes

### Stability Analysis

In order to check the model’s stability, a set of individual transformations were done on the input images and the model’s results for these transformed images were compared to the results of the original image. The transformations were as follows:

A. Contrast changes.
B. Rotation by 5 degrees.
C. Scaling.
D. Translation by a factor.
E. Adding a gaussian noise.

An example of these transformations can be found in **Figure 2**.

**Figure 2:**
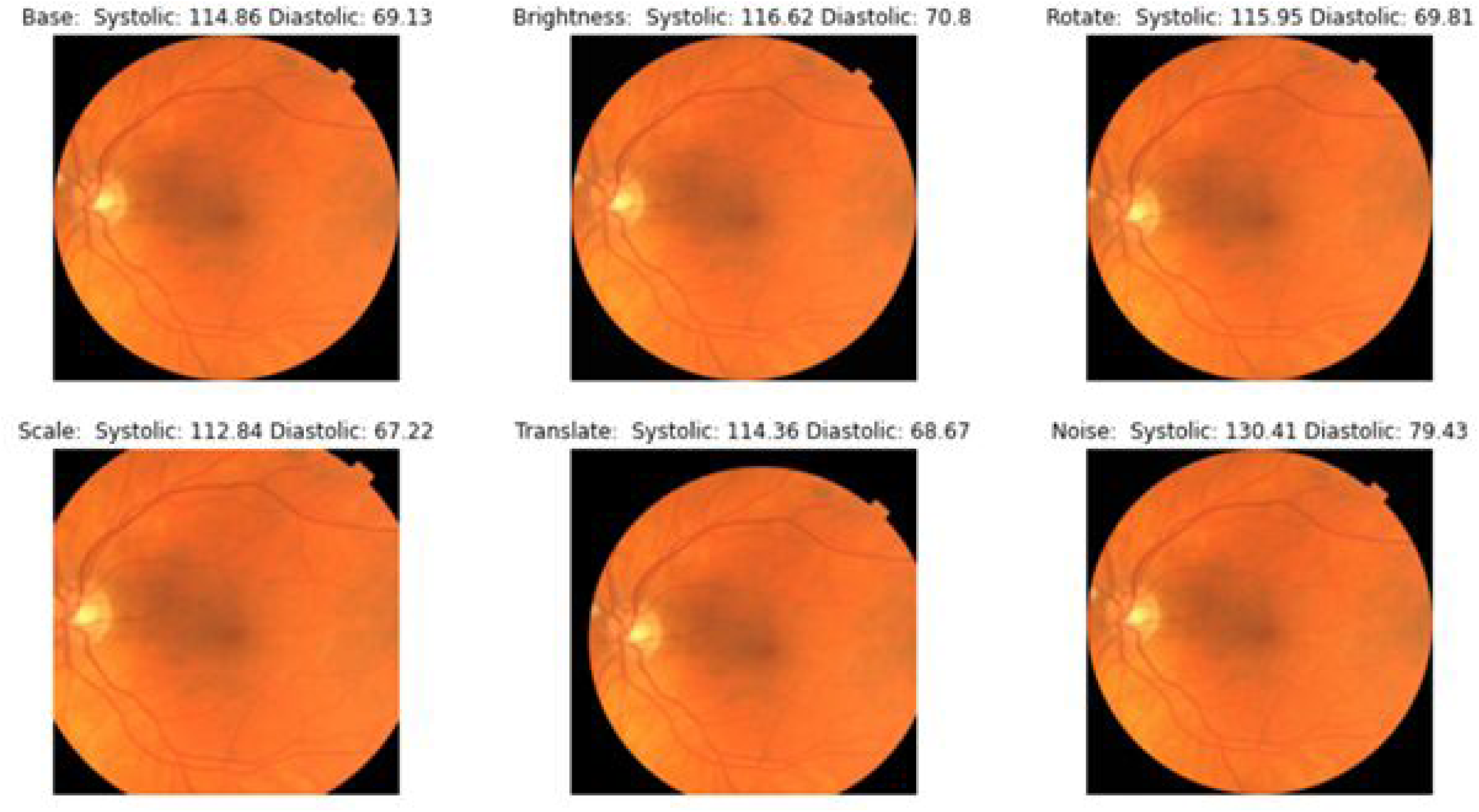
Example of stability check results for one left eye following transformations

Comparison was done by calculating the mean absolute errors between the original image and the transformed image, and finally calculating the average and standard deviation of errors for each transformation.

## Results

**Table 2** presents the model result on the test dataset with comparison to prior state of the art results. Systolic BP was measured with 9.81 (CI 95% 6.97, 10.01) MAE, 165.13 (CI 95% 156.44, 174.70) MSE and 0.36 (CI 95% 0.34, 0.38) R^2. Diastolic BP was measured with 6.00 (CI 95% 5.86, 6.14) MAE, 58.21 (CI 95% 55.62, 61.06) and 0.30 (CI 95% 0.28, 0.33) R^2. Additional stratifications of model results across sex, ethnicity, and age, in terms of MAE, MSE, and R^2, are shown in supplementary tables X-Y.

**Table 2:** BP model results (95% CI) compared to Poplin et al^25^.

|  | MAE | MSE | R<sup>2</sup> |
| --- | --- | --- | --- |
| Systolic | 9.81 (9.67, 10.01) | 165.13 (156.44, 174.70) | 0.36 (0.34, 0.38) |
| Diastolic | 6.00 (5.86, 6.14) | 58.21 (55.62, 61.06) | 0.30 (0.28, 0.33) |
| Systolic, Poplin et al | 11.35 (11.18, 11.51) | N/A | 0.36 (0.35, 0.37) |
| Diastolic, Poplin et al | 6.42 (6.33, 6.52) | N/A | 0.32 (0.30, 0.33) |

Figure 1 shows the model’s focus areas using Grad-CAM, demonstrating areas of attention around the optic disc and major blood vessels.

**Table 3** shows the results for the stability analysis in terms of mean absolute error for each transformation displayed in Figure 2, showing high stability in morphological transformations and slightly lower stability in color and noise transformations.

**Table 3:** Stability analysis results.

|  | Systolic Absolute Error Mean (S.D) | Diastolic Absolute Error Mean (S.D) |
| --- | --- | --- |
| Brightness | 7.45 (5.70) | 4.10 (3.01) |
| Rotation | 1.68 (1.34) | 1.00 (0.83) |
| Scale | 2.91 (2.27) | 2.12 (1.51) |
| Translation | 1.96 (1.44) | 1.20 (0.90) |
| Noise | 8.22 (5.74) | 4.46 (3.10) |

Although the UK BioBank data included consists of ∼84% British citizens, the vast majority of whom are Caucasian, results are comparable across different ethnicities (see **supplementary tables 3-4**); given the large size of database, statistically meaningful results were obtained from other demographics as well.

## Discussion

This work introduces a novel DL method to estimate instantaneous systolic and diastolic BP from a single fundus image. Recent work by Poplin et al.^22^ was able to estimate systolic BP according to fundus images with an acceptable error rate. While the algorithms predicted systolic blood pressure (SBP), BMI, and HbA1c better than baseline, these predictions had low coefficient of determination suggesting that the algorithm is not able to predict these parameters with high precision. The predicted SBP increased linearly with actual SBP until approximately 150 mmHg but levelled off above that value.

Our model analyzes macroscopic anatomic features of the retina as they appear in a fundus image to assess BP. Data from Grad-CAM images suggest that the model relies mostly on blood vessel architecture and optic disc morphology. The current work shows significant improvement over Poplin’s results (see Table 2), demonstrating the relative stability of BP measurement via fundus images.

Estimating the true error rate of cuff measurements is difficult, even as it is the gold standard and most widely used method to assess BP. A previous study by Eilertsen and Humerfelt found the average values of BP recorded by trained individual observers to vary by as much as 5 to 10 mmHg.^29^ An additional meta-analysis aimed to compare the accuracy of cuff measurement to intra-arterial measurements, finding a mean absolute difference of 8mmHg between cuff and intra-aortic measurement,^30^ with cuff BP overestimating diastolic intra-arterial measured BP by 5.5 mmHg. The accuracy of cuff measurement was best for normal BP and stage 2 hypertension and above, and worst for pre-hypertension and stage 1 hypertension.

Additionally, multiple factors can greatly influence any single cuff measurement. For example, engaging in conversation while measuring the pressure raises the systolic and diastolic BP by 10 mmHg.^31^ A distended bladder may raise BP by >10 mmHg, sitting with the arm unsupported may raise the diastolic pressure by 10 mm Hg, and ill-fitting cuff size, wearing the cuff over clothing, or smoking before the exam may also have effect.^32^ White coat hypertension is another well-known pitfall, shown to raise BP by up to 25 mmHg^13,31^ and affecting 15%-25% of individuals. BP readings also follow a diurnal pattern, potentially leaving long nocturnal periods of hypertension unmeasured and unaccounted for during regular outpatient office visits.

The results of our model revealed a mean absolute error (MAE) of 9.8 mmHg, which is comparable to the intra patient varies of the cuff measurement. Of note is the fact that in our model the ground truth for BP was the cuff measurement, which was taken in a standardized fashion by trained personnel,^26^ but has its own error rate, as mentioned. We estimate that our model will be uninfluenced by the above-mentioned pitfalls, as it relies on anatomic factors which are not as easily manipulated and labile.

Due to the sometimes-unpredictable nature of cuff BP measurements, and the multiple causes for variance in measurement, ambulatory BP monitoring (ABPM) has been utilized for more accurately assessing BP.^33^ This is achieved by equipping the patient with a wearable device which measures BP in the patient’s natural environment, usually for a period of 24 hours. This method has been shown in multiple studies to provide a better estimate of the risk of cardiovascular disease than single office BP measurement, but the approach is limited by cost, availability, and patient comfort.^34,35^

The effects of hypertension on the retina are well documented and occur in three stages. The vasoconstrictive stage is caused by vasospasm and autoregulation in the arteriole, directed at modulating flow through the arteriole and evident as generalized narrowing. The sclerotic phase occurs as a result of arteriosclerosis in the retina, causing intimal thickening, hyperplasia of the middle wall, and hyaline degeneration. The manifestations are severe generalized and focal areas of arteriolar narrowing, AV nicking, and vessel wall opacification (otherwise termed silver or copper wiring). In the exudative stage, severe hypertension disturbs the blood retinal barrier, causing leakage of blood and lipids into the retina, ischemia, and possibly disc swelling.^14,15,36^ Most of these morphologic features are related to chronic hypertension. AV nicking and generalized arteriolar narrowing were specifically found to be associated with chronic hypertension regardless of current BP measurement.^16,17^

It is important to note that our model, as well as the previous model published by Poplin,^22^ aims to estimate the BP in individuals without overt hypertensive retinopathy, or a previous diagnosis of hypertensive retinopathy. As shown, our model focuses greatly on blood vessels when making the assessment of BP. We hypothesize that our algorithm recognizes changes in retinal arterial caliber and composure which may be subtle and easily missed by the ophthalmologist. The Modified Scheie Classification of Hypertensive Retinopathy^37^ is an accepted classification scheme of hypertensive retinopathy. In this system, grade 1 hypertensive retinopathy is classified as barely detectable arterial narrowing. This supports the notion that early retinal changes in this setting may be subtle and easily overlooked. A Lancet publication also concluded that early retinopathy grades may be difficult to distinguish by ophthalmoscopy.^38^

We therefore hypothesize that our model may provide a better representation of chronic BP status over time rather than the acute nature of a single BP measurement, which is also subject to multiple biases. We believe that our model may provide a more balanced and stable BP assessment, much like an ABPM can provide an average measurement compared to a single BP measurement. The BP value generated by our model may represent BP values for the specific patient over a period of months to years, but further studies will be needed to verify this hypothesis. We suggest that utilizing our model instead of taking a single cuff measurement may be analogous to using a hemoglobin A1C result as compared to a single glucose value for monitoring a diabetic patient.

In this work, any patient with a record of BP medication use was excluded from the analysis. The rationale was to avoid a disconnect between the cuff BP measurement, which may be temporarily reduced by medication, and the status of the retinal vasculature which represents a more prolonged BP effect as discussed.^16,17^ In addition, BP therapies may alter ocular structures and influence disease risk including glaucoma. Specifically, a large population-based study found persons free from eye disease undertaking all classes of antihypertensive medications were associated with larger cup size and higher C/D ratio in subjects with either DBP<90 mm Hg or SBP<140 mm Hg.^39^ Going forward, accounting for BP therapeutics will be important to understand impact of treatment and potentially provide novel targets for assessing benefit.

Our work has some limitations including that the longitudinal impact of modeled BP to disease outcomes was not assessed. Additionally, the mean age of the sample was ∼56 years of age, limiting the ability to include the impact of aging on imagery and DL outcomes. The exclusion of persons under BP reducing therapies also limits applicability to persons under medical management for hypertension.

Future studies should include cohorts under hypertensive therapies to assess the long-term benefits or risk of treatments on the vasculature. For instance, several pilot studies were able to show that antihypertensive medication can actually reverse some of the microvascular changes in hypertensive retinopathy.^40,41^ Specifically, one study found Enalapril reduced retinal arterial wall opacification after 26 weeks of treatment.^41^ These effects, however, were true for only some medications and only a part of the changes caused by hypertensive retinopathy. It is unclear what the impact of these medications would be on the retinal vascular morphology of eyes without a diagnosis of overt hypertensive retinopathy.

The significant and rising impact of hypertension and associated disease globally necessitates improved diagnostics for providing individualized precision care. Ideally novel targets would be non-invasive, highly accessible, and have limited error and provider introduced variability. In addition, improved BP assessment should better represent the chronic condition and provide insight on microvasculature functionality rather being a snapshot of arm pressure. In this analysis we demonstrate the ability to predict systemic BP from fundus photographs with high specificity and acceptable error compared to cuff measurements. Our approach has the benefit of providing a better understanding of chronic versus acute status of BP due to the long-term effects on the ocular vasculature and reduced operator error and white coat effects. Long-term studies are needed to understand the impact of these biomarkers on disease status.

## Data Availability

This research has been conducted using the UK Biobank Resource under Application Number 75750. UK Biobank has been established as an open-access resource for public health research, with the intention of making the data as widely available as possible in an equitable and transparent manner.

https://www.ukbiobank.ac.uk/

## Acknowledgements and Funding

ZDA and IB contributed research design. ZDA, IB, and RA participated in data acquisition and/or research execution. ZDA and IB contributed data analysis. IB, DD, AH, BS, RA, DM, and TI prepared the manuscript. ZDA and IB had access to study data.

Employees and board members of AEYE Health designed and carried out the study; managed, analysed and interpreted the data; prepared, reviewed, and approved the article; and were involved in the decision to submit the article.

